# Comparative Analysis of Task-Specific and Combined Upper-Limb EMG Features for Early Parkinson’s Disease Classification

**DOI:** 10.64898/2026.03.16.26348216

**Authors:** Júlia Rey Vilches, Chiara Gorlini, Silvia Tolu, Trine Hørmann Thomsen, Bo Biering-Sørensen, Sadasivan Puthusserypady

**Author notes:** Emails: {;,;,;}, }.

## Abstract

Early-stage Parkinson’s disease (PD) presents motor impairments that are difficult to detect clinically. Surface EMG (sEMG) offers an objective alternative, yet many studies rely on non-standardized tasks and provide limited task- and symptom-specific interpretability. This study analyzes sEMG recorded during standardized MDS-UPDRS-III upper-limb tasks—pronation–supination and postural tremor—performed by individuals with early-stage PD (Hoehn and Yahr 1–3, n=31) and healthy controls (n=30). Time-, frequency-, and nonlinear features were extracted and evaluated using a two-stage framework combining filter-based ranking and wrapper-based methods to support feature selection across multiple classifiers and interpretability. Pronation–supination showed the strongest single-task discrimination (balanced accuracy 0.79±0.181), driven by rhythm and nonlinear features reflecting impaired rhythmicity, reduced neuromuscular complexity, and unstable muscle deactivation, consistent with bradykinesia and rigidity. The postural tremor task highlighted tremor-specific spectral changes and reduced signal complexity during sustained posture (balanced accuracy 0.75±0.18), capturing low-frequency oscillations typical of PD tremor. Combining both tasks further improved classification without increasing feature dimensionality (balanced accuracy 0.83±0.186), indicating complementary diagnostic information. Filter-guided selection enhanced robustness and consistency across models. Beyond classification, this study highlights the value of interpretable, task-aligned motor quantification, showing that standardized clinical movements combined with targeted sEMG analysis can support explainable assessment of early-stage PD motor symptoms.

## I. Introduction

Parkinson’s disease (PD) is a progressive neurodegenerative disorder that manifests through both motor and non-motor symptoms, progressively reducing autonomy and quality of life [1]. Motor symptoms play a central role in both the patient experience and clinical diagnosis. Bradykinesia, rigidity, and tremor are among the key motor features of PD and are typically the earliest manifestations that prompt clinical evaluation [2]. These motor symptoms often appear gradually, asymmetrically, and can fluctuate over time, making early-stage PD particularly difficult to identify with confidence. As a result, clinical diagnosis still relies primarily on expert neurological assessment, and even experienced movement disorder specialists misdiagnose early or atypical cases in up to 30% of instances, with diagnostic accuracy further reduced among general neurologists [3] [4].

The Movement Disorder Society Unified Parkinson’s Disease Rating Scale, Part III (MDS-UPDRS-III), is the standard clinical instrument for assessing motor symptoms in PD [5]. It comprises a set of well-defined, standardized motor tasks designed to probe different motor domains, including bradykinesia, rigidity, and tremor. However, despite its widespread use, its scoring remains subjective and is influenced by inter-clinician variability, and at the same time the test is highly time-consuming in clinical practice [3].

In response to these limitations, there has been growing interest over the past decades in the development of objective and quantifiable indicators of PD [6]. Wearable sensors enable the analysis of motor performance with increased temporal and spatial resolution [7] [8]. Among these, surface electromyography (sEMG), a well-established technique in neuromuscular research, captures muscle activation patterns and motor unit behavior, providing direct insight into neuromuscular function [9] [10].

In practice, the application of sEMG in PD assessment has remained fragmented. Most studies focus on tremor, characterizing low-frequency oscillatory activity from antagonistic forearm muscles using frequency-domain features [11], [12]. sEMG-based PD discrimination or motor score prediction has also been explored using machine-learning approaches, but these often rely on non-standardized tapping tasks, non-task-specific muscle placement, or cohorts biased toward mid-to advanced-stage disease, limiting task and symptom-specific interpretability [13], [14].

Beyond conventional time and frequency domain descriptors, evidence of altered muscle coordination and reduced signal complexity in PD highlights the potential relevance of multi-domain sEMG features, including nonlinear metrics [1], [15], [16]. However, such features are rarely investigated within standardized clinical tasks or evaluated in a task and muscle-specific framework, constraining their clinical interpretability and translational value.

In this work, we aim to address these gaps through a comparative, task-specific analysis of sEMG features extracted from standardized MDS-UPDRS-III upper-limb tasks in an early-stage PD cohort. Two clinically relevant tasks were selected: pronation–supination of the hands, representing cyclic voluntary movement, and postural tremor assessment, representing sustained postural activation. These tasks test complementary motor control mechanisms that are routinely evaluated in clinical practice.

A comprehensive set of sEMG features spanning time-domain, frequency-domain, and nonlinear descriptors is extracted and analyzed within a two-stage framework designed to support both classification and interpretability. This framework enables a systematic comparison of task-specific and combined feature sets, allowing investigation of which features and feature domains are most informative, how discriminative performance differs across standardized motor tasks, and whether combining tasks provides complementary diagnostic information. Through this approach, the study seeks not only to distinguish PD from healthy controls, but also to clarify how standardized movements and specific sEMG feature domains contribute to the quantification of early PD motor impairment.

## II. Study design and methods

The study analyzed sEMG data collected from individuals with early-stage PD (Hoehn and Yahr stages [17], H&Y, 1–3, n=31) and HC (n=30) during upper-limb motor tasks routinely performed as a part of the MDS-UPDRS-III clinical motor examination [5]. All recordings followed a previously published and fully standardized acquisition protocol [18]. Participant demographic characteristics are summarized in Table I.

**TABLE I.** Participant demographics and clinical characteristics.

| Group | N | Age (years) | Range | Sex (F/M) | H&Y (1/1.5/2/3) |
| --- | --- | --- | --- | --- | --- |
| PD | 31 | $62.6 \pm 9.1$ | 43–79 | 14/17 | 17/4/7/3 |
| HC | 30 | $59.2 \pm 13.6$ | 40–79 | 18/12 | – |

To assess distinct aspects of upper-limb motor impairment in PD, two tasks from the MDS-UPDRS-III were analyzed (Figure 1). Table II summarizes the selected tasks, including their clinical purpose, movement characteristics, and corresponding sEMG recordings.

**Fig. 1:**
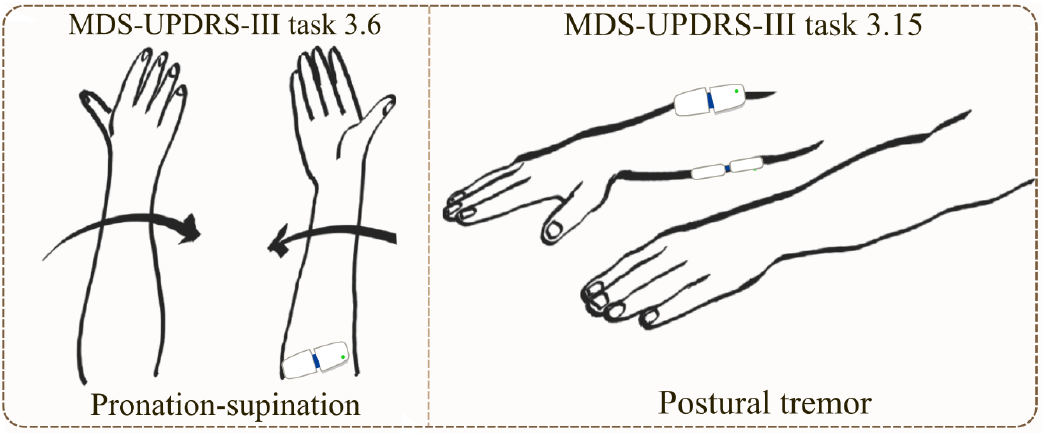
Upper-limb motor tasks analyzed from the MDS-UPDRS-III, with corresponding sensor placement.

**TABLE II.** Overview of analyzed MDS-UPDRS-III upper-limb motor tasks.

| Task | Clinical task | Movement description | sEMG muscles | Motor features assessed |
| --- | --- | --- | --- | --- |
| 3.6 | Pronation–supination | Rapid alternating pronation and supination of the hands with arms extended | Pronator Teres (PT) | Bradykinesia-related features: movement speed, amplitude, hesitations and halts |
| 3.15 | Postural tremor | Arms extended in front with palms facing downward, maintaining a static posture | Flexor Carpi Radialis (FCR)<br>Extensor Carpi Radialis (ECR) | Sustained muscle activation and oscillatory tremor activity |

Together, they capture both cyclic movements where bradykinesia can be detected and sustained postural activation where rigidity- and tremor-related features may emerge. sEMG signals were recorded unilaterally from one upper limb, corresponding to the symptom-dominant side in participants with PD. Muscle activity was acquired using MuscleBAN BLE surface EMG sensors (BioPlux).

### A. Signal processing

Raw sEMG signals were sampled at 1000 Hz and processed using a standardized pipeline implemented in Python. Each channel was first demeaned to remove DC offset. Signals were then band-pass filtered using a fourth-order Butterworth filter [19] [20] with cutoff frequencies of 1–450 Hz to suppress motion artifacts and high-frequency noise outside the physiological EMG bandwidth, while preserving low-frequency components associated with tremor activity [21] [22] [23]. To reduce power-line contamination, a second-order IIR notch filter centered at 50 Hz was applied to the band-passed signal [24]. After processing, both the band-pass-filtered signal and a full-wave rectified version obtained by taking the absolute value were retained for subsequent analyses.

For the pronation-supination task (Task 3.6), an sEMG envelope was computed for segmentation purposes by rectifying the filtered signal, and applying a low-pass fourth-order Butterworth filter (10 Hz cutoff). Contraction onsets and offsets were identified using a double-threshold method with thresholds set at the 45th and 60th percentiles of the envelope, and relaxation intervals were defined as inter-burst periods (Figure 2). The first and last 3 seconds of the recording were discarded to remove transient effects, and analyses were performed on both the full signal and the segmented contraction and relaxation intervals [25]. For the postural tremor task (Task 3.15), no segmentation was applied. After processing (demeaning, band-pass filtering, notch filtering, and rectification), the first and last 3 seconds of the recording were cropped to minimize transient effects. Subsequent analyses were conducted on the remaining continuous signal.

**Fig. 2:**
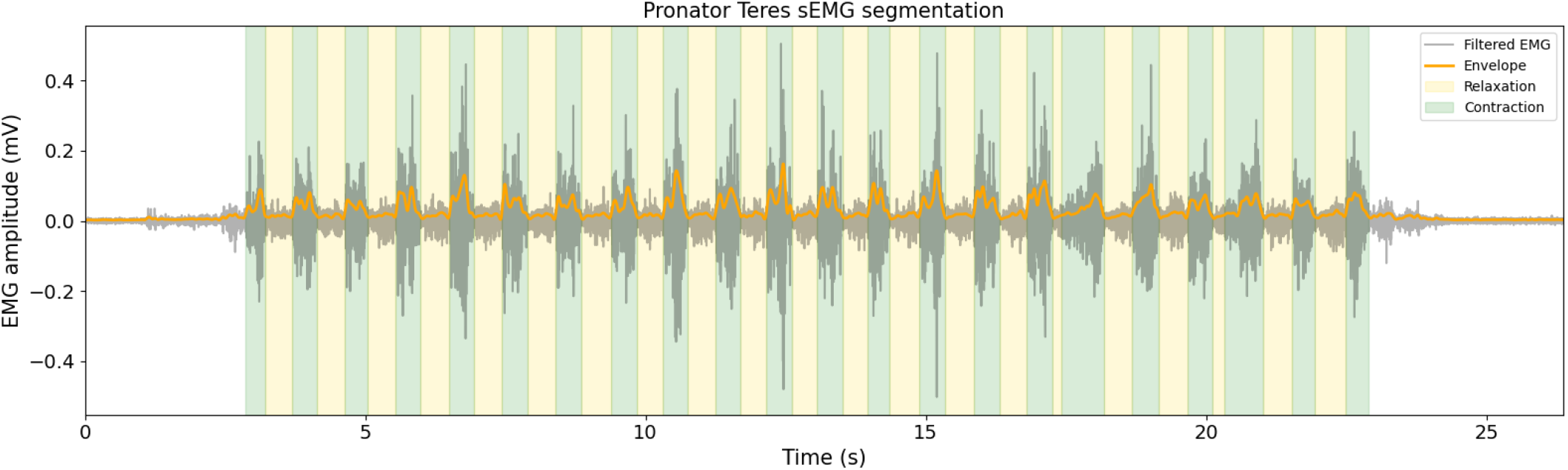
Example of sEMG signal segmentation for Task 3.6 (pronation–supination). The filtered signal, rectified envelope, and detected contraction and relaxation intervals are shown.

### B. Feature extraction

Feature extraction was performed separately for each participant, task, and muscle, computing a multi-domain set of time, frequency, and nonlinear features. For Task 3.6, features were extracted from the full signal, and from contraction and relaxation segments. Segment-level features were summarized across bursts using the mean, standard deviation, minimum, maximum, and median. For Task 3.15, features were computed from the continuous, unsegmented signal. From the full filtered signal, time-domain features included root mean square (RMS), variance, mean absolute value, and zero crossings. Frequency-domain features, derived using Welch’s power spectral density, included total power, mean and median frequency, tremor-band power (2–6 Hz), tremor-to-total power ratio, and band-limited power in the low (5–40 Hz), mid (40–90 Hz), and high (90–250 Hz) frequency bands [26]. Nonlinear features, describing signal complexity, regularity, and dynamical stability, included the Lyapunov exponent, Higuchi fractal dimension, and sample entropy [1].

From the full rectified signal, rhythm-related features included the mean inter-contraction interval (ICI; time between successive contraction peaks) and the mean inter-burst interval (IBI; time between activation bursts). In Task 3.6, additional time-domain features were computed from the contraction and relaxation segments, including slope sign changes (SSC), integrated EMG (iEMG) [27], peak amplitude, and segment duration. Specifically, for contraction segments, envelope-derived rise and fall times were also computed. In Task 3.15, inter-muscle coordination between the two recorded muscles was assessed using envelope-derived Pearson correlation, and the RMS ratio between the muscles.

### C. Statistical analysis and filter-based feature ranking

A total of 261 sEMG features were extracted across tasks, muscles, and feature domains. Given this high-dimensional feature space, an initial feature-ranking step was required to assess the relative importance of individual features with respect to disease status and to guide subsequent multivariate modeling.

As a first step, filter-based feature ranking was applied to evaluate the relationship between each feature and class membership (PD vs HC) independently [28]. For each task and muscle, features were analyzed separately. Group-level differences were assessed using univariate statistical tests, including independent two-sample t-tests and one-way analysis of variance (ANOVA), to evaluate differences in feature means between groups. In parallel, Pearson’s correlation coefficient was computed to quantify linear associations between individual features and the binary class label. To account for potential nonlinear dependencies not captured by linear statistics, mutual information between each feature and the class label was additionally computed. Together, these complementary metrics provided an initial, model-agnostic assessment of feature relevance across the full feature set [29].

Based on these relevance scores, features were ranked per task, and the top-ranked features were retained to form a reduced candidate feature pool, limiting the number of features to fewer than 30 per task. This step aimed to reduce redundancy, improve interpretability, and constrain the search space for subsequent modeling. In a second step, wrapper-based feature selection was applied to evaluate feature interactions and determine multivariate feature importance in the context of classification [30] [31]. Wrapper methods iteratively trained and evaluated classification models using different feature subsets, selecting combinations that maximized classification performance between PD and HC groups.

To assess whether the filter-based ranking step was beneficial and necessary, the wrapper-based selection was performed both on the reduced, top-ranked feature pool and on the full feature set, allowing direct comparison of performance and feature selection behavior between the two approaches.

### D. Wrapper-based feature selection and classification

Following filter-based ranking, a wrapper-based feature selection strategy was used to identify compact, task-specific, and joint feature subsets that maximize classification performance. In addition to optimizing classification, this step was designed to compare the relative discriminative contribution of individual motor tasks and to assess whether combining feature sets across tasks provides complementary information beyond single-task analysis. This enables evaluation of task-specific relevance as well as the added value of multi-task feature integration for PD classification.

Wrapper-based selection was performed separately for task 3.6, task 3.15, and a joint feature set, enabling direct comparison of task-specific and combined-task feature subsets. An exhaustive subset search (ESS) strategy was employed to evaluate all possible feature combinations with subset sizes ranging from 1 to 10.

Five classification models were evaluated: logistic regression (LR), support vector machine with linear kernel (SVM-linear), support vector machine with radial basis function kernel (SVM-RBF), random forest (RF), and k-nearest neighbors (k-NN). For each feature subset and classifier, a standardized pipeline consisting of z-score normalization followed by model fitting was applied. Classification performance was estimated using stratified 10-fold cross-validation to preserve class balance across folds. The classifiers were selected as complementary, well-established methods spanning linear and nonlinear, parametric and non-parametric models [32].

Model performance was primarily quantified using balanced accuracy, defined as the mean of sensitivity and specificity (Eq. 1). Receiver operating characteristic area under the curve (ROC–AUC) was additionally computed as the area under the ROC curve obtained by varying the decision threshold, providing a threshold-independent measure of class separability [33]. For each task set and classifier, feature subsets were ranked according to mean cross-validated balanced accuracy, with ROC–AUC used as a secondary criterion.

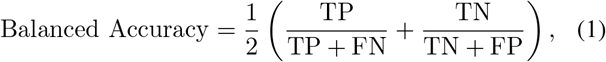

where TP and TN denote the number of true positives and true negatives, respectively, and FP and FN denote false positives and false negatives.

## III. RESULTS

### A. Feature relevance across standardized motor tasks

Filter-based feature ranking was used to obtain an initial assessment of feature relevance prior to multivariate modeling. For Task 3.6, highly ranked features predominantly reflected rhythm-related and nonlinear characteristics of the PT, including variability in relaxation duration and fall time, slope sign changes, inter-burst timing, and Higuchi fractal dimension. In contrast, for Task 3.15, top-ranked features were largely drawn from the frequency and nonlinear domains, particularly mean and median frequency, tremor-band power, sample entropy, and Higuchi fractal dimension derived from the ECR and with more dominance FCR muscles (Figure 3). Across both tasks, nonlinear features consistently appeared among the most relevant descriptors, alongside selected frequency-domain metrics, whereas amplitude-based time-domain features were less prominent.

**Fig. 3:**
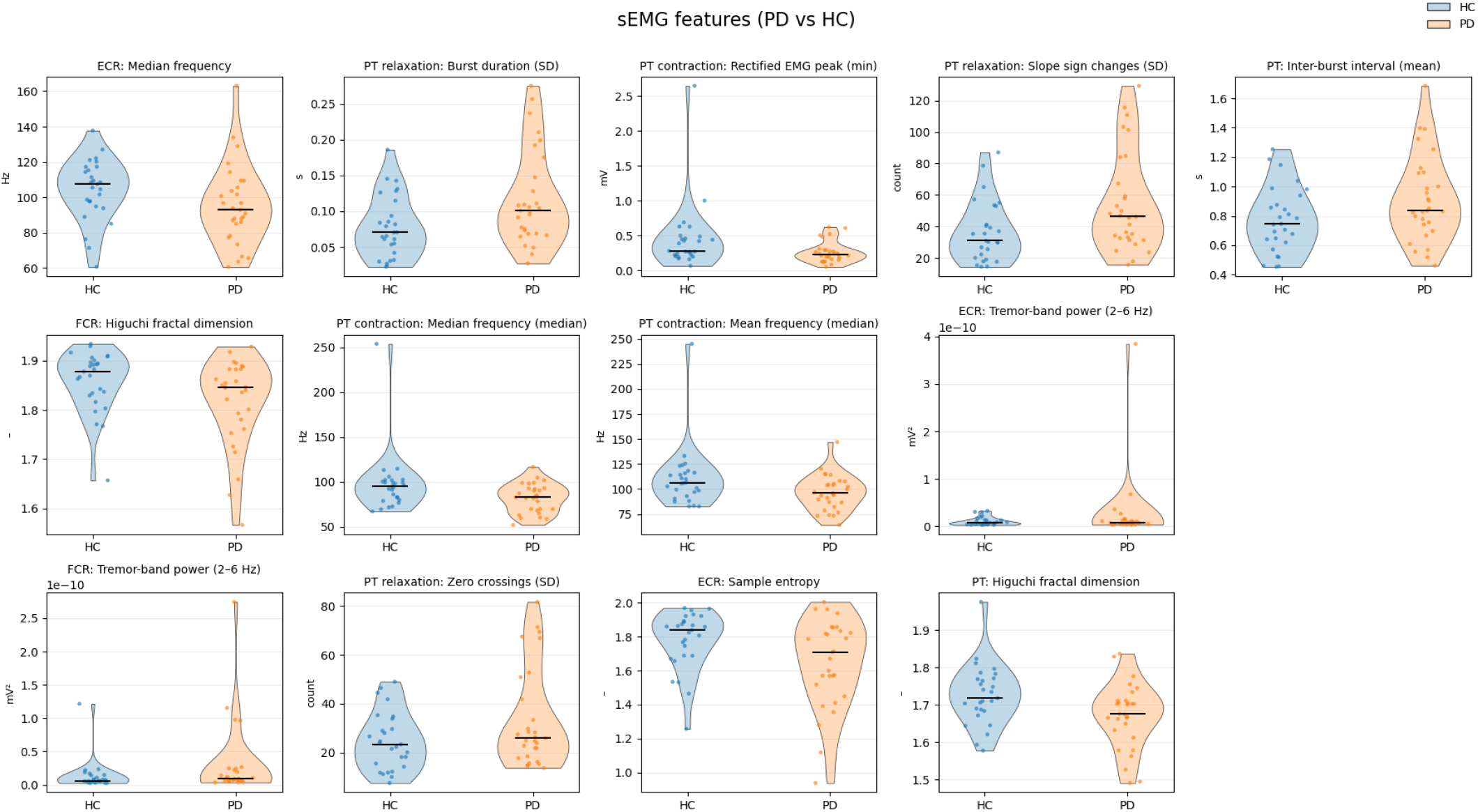
Group-level distributions of representative sEMG features across the analyzed MDS-UPDRS-III tasks. Violin plots compare early-stage PD and HC participants, illustrating task-specific differences across selected sEMG feature domains. Features without physical units are indicated by a dash (“–”) on the x-axis.

### B. Task-specific classification performance

Wrapper-based feature selection and classification were performed separately for task 3.6 and task 3.15. Classification performance for all evaluated models is summarized in Table III.

**TABLE III.** Classification performance for task-specific (Task 3.6, Task 3.15) and joint sEMG feature sets. Balanced accuracy and ROC–AUC are reported as mean ± standard deviation across cross-validation folds. #Feat. denotes the number of selected sEMG features used by the corresponding model.

| Task | Model | Bal. Acc. | ROC-AUC | #Feat. |
| --- | --- | --- | --- | --- |
| 3.6 | SVM-lin | **0.792 ± 0.181** | 0.772 ± 0.213 | 5 |
| | RF | 0.775 ± 0.162 | 0.706 ± 0.262 | 4 |
| | KNN | 0.758 ± 0.169 | 0.769 ± 0.193 | 5 |
| | SVM-RBF | 0.750 ± 0.124 | 0.756 ± 0.193 | 4 |
| | LR | 0.733 ± 0.175 | 0.739 ± 0.196 | 5 |
| 3.15 | SVM-RBF | **0.750 ± 0.180** | 0.744 ± 0.235 | 4 |
| | SVM-lin | 0.742 ± 0.159 | 0.750 ± 0.215 | 2 |
| | LR | 0.725 ± 0.136 | 0.711 ± 0.185 | 2 |
| | KNN | 0.708 ± 0.189 | 0.703 ± 0.244 | 3 |
| | RF | 0.700 ± 0.233 | 0.681 ± 0.137 | 5 |
| Joint | KNN | **0.825 ± 0.186** | 0.789 ± 0.246 | 5 |
| | SVM-lin | 0.817 ± 0.183 | **0.844 ± 0.230** | 4 |
| | LR | 0.800 ± 0.205 | 0.828 ± 0.223 | 5 |
| | SVM-RBF | 0.792 ± 0.132 | 0.711 ± 0.247 | 4 |
| | RF | 0.775 ± 0.184 | 0.767 ± 0.243 | 3 |

For Task 3.6, the highest balanced accuracy was achieved using a SVM-linear classifier, yielding a balanced accuracy of 0.792± 0.181 and a ROC–AUC of 0.772± 0.213 selecting a subset of five features. For task 3.15, the best-performing model was an SVM-rbf, achieving a balanced accuracy of 0.750± 0.180 and a ROC–AUC of 0.744 ±0.235 using four features (Table III). Classification performance exhibited notable variability across folds, suggesting the presence of overlapping feature distributions between early PD and HC participants.

### C. Comparison of single-task and joint-task feature sets

To assess whether combining standardized motor tasks provides complementary information, wrapper-based selection was repeated using a joint feature set comprising features from both already analyzed tasks. Results are reported in Table III. The joint-task configuration yielded the highest overall classification performance across all task sets. Using a KNN classifier, a balanced accuracy of 0.825± 0.186 and a ROC–AUC of 0.789± 0.246 were achieved with five selected features. Comparable performance was observed for SVM-linear and LR models. Across classifiers, the joint-task feature set consistently improved balanced accuracy compared to single-task analyses. This improvement was achieved without increasing the number of selected features, indicating that combining cyclic movement (3.6) and sustained postural (3.15) tasks expands the discriminative feature space while maintaining compact feature subsets.

### D. Effect of filter-based feature ranking

To evaluate the impact of filter-based feature ranking, wrapper-based classification was additionally performed without prior filtering. A comparison of the best-performing models and selected feature subsets with and without filtering is shown in Table IV.

**TABLE IV.**
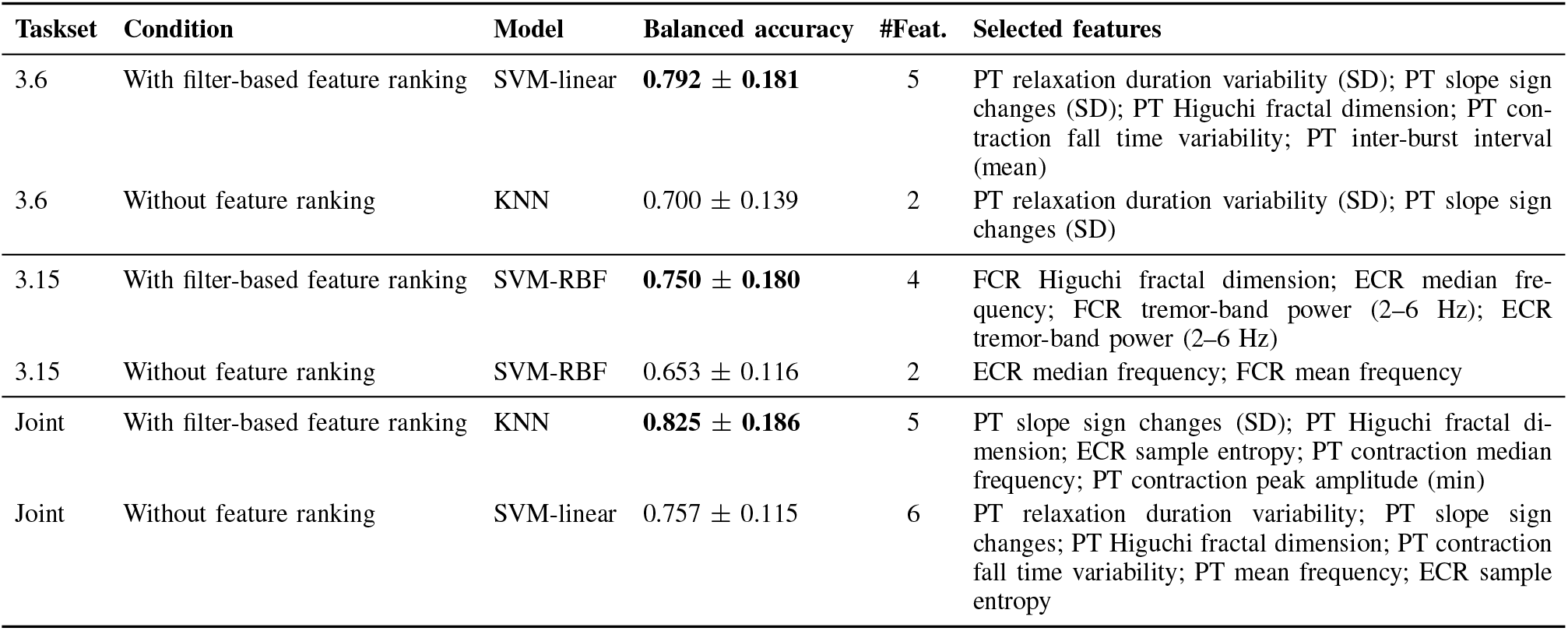
Comparison of best-performing wrapper-based models with and without prior filter-based feature ranking.

Across all task configurations, classification performance was consistently reduced when wrapper-based selection was applied directly to the full feature set without filtering. In addition to lower balanced accuracy, unfiltered analyses generally selected fewer features with reduced performance.

Despite these differences, several key features were consistently selected with and without filtering, notably rhythm-related and nonlinear descriptors for task 3.6 and frequency-domain features for task 3.15. Working with filter-based ranked features, therefore reducing the feature set, improved robustness in the performance of the wrapper-based classification.

### E. Selected feature subsets and interpretability

Wrapper-based feature selection provided compact and interpretable feature subsets across task-specific and joint-task analyses. Figure 3 shows the distributions of representative features selected for discrimination between PD and HC.

For task 3.6 (pronation–supination), most selected features reflected movement timing, rhythm, and signal complexity. In PD, rhythm-related features—including relaxation duration variability, inter-burst interval, and slope sign changes—were elevated, indicating increased irregularity and impaired contraction–relaxation transitions consistent with rigidity and bradykinesia. PT-derived frequency-domain features (mean and median frequency) were generally lower, suggesting slower motor unit recruitment, while reduced Higuchi fractal dimension and sample entropy indicated diminished neuro-muscular complexity during cyclic movement.

For task 3.15 (postural tremor), selected features that were derived from both FCR and ECR emphasized tremor-related spectral content. Tremor-band power was consistently higher in PD for both muscles, with a more pronounced separation observed in the FCR, reflecting increased oscillatory activity during sustained postural activation. Median frequency values, particularly for the ECR, were lower in PD, indicating a dominance of low-frequency components during static muscle activation, proving a difference in muscle behavior during sustained muscle engagement.

Nonlinear features, including Higuchi fractal dimension and sample entropy, also contributed to class separability task 3.15. Lower values in PD indicate more regular and less complex muscle activation patterns, simiarly to the case even in dynamic movements (3.6).

Joint-task analysis showed that rhythm and nonlinear features from task 3.6, along with complexity features from task 3.15, were most effective for classification. PT-derived features dominated, but nonlinear ECR features added complementary information on postural tremor control.

## IV. Discussion and conclusion

This study examined the discriminative and interpretative value of sEMG features extracted from standardized MDS-UPDRS-III upper-limb tasks in individuals with early-stage PD, with emphasis on task specificity and feature-domain contributions. Focusing on well-defined motor tasks helps mitigate known limitations of EMG-based assessment related to symptom evaluation outside controlled or task-specific conditions [34].

Across all analyses, the pronation–supination task (Task 3.6) consistently emerged as the most informative. Features originating from the pronator teres muscle—especially rhythm-related and nonlinear descriptors—were repeatedly selected in the highest-performing models. Increased variability in contraction–relaxation timing and more frequent slope sign changes in PD participants indicated impaired rhythmic motor control and unstable muscle deactivation, aligning with bradykinesia- and rigidity-related dysfunction. Reductions in frequency-domain metrics and nonlinear complexity further suggested slower motor unit recruitment and diminished adaptability during voluntary cyclic movement.

In contrast, the postural tremor task (Task 3.15) was dominated by tremor-related spectral features and reduced signal complexity in the forearm flexor and extensor muscles. Elevated tremor-band power and lower median frequency values reflected increased low-frequency oscillatory activity during sustained posture, while reduced entropy and fractal dimension indicated more regular, synchronized muscle activation patterns characteristic of PD tremor.

Recent sEMG studies have used standardized MDS-UPDRS-III upper-limb tasks to investigate task-dependent muscle activation patterns. For example, [12] showed that different UPDRS tasks present different low-frequency sEMG signatures, supporting the idea of task specificity. However, their analysis was conducted only on healthy adults and did not account for task-relevant muscles, as all movements were recorded from the same antagonistic muscle pair, without addressing disease discrimination. Similarly, [35] demonstrated that simple time-domain sEMG features extracted from rhythmic MDS-UPDRS-III hand movements can capture aspects of bradykinesia in PD. However, their analysis focused exclusively on one motor symptom, cyclic tasks, relied on a narrow feature set, and involved a small cohort that did not include individuals in the early stages of the disease.

Building on these foundations, the present study focuses on early-stage PD and combines complementary MDS-UPDRS-III upper-limb tasks with a multi-domain, muscle-specific sEMG analysis. This enables a systematic comparison of cyclic and sustained motor tasks and provides a more comprehensive, interpretable characterization of distinct motor impairments.

Combining feature sets from both tasks produced the highest classification performance without increasing feature dimensionality, demonstrating that voluntary cyclic and sustained postural tasks provide complementary diagnostic information. The predominance of Task 3.6 features in joint-task subsets underscores the importance of rhythmic voluntary control in early PD, while Task 3.15 contributed additional tremor-related insights.

Finally, comparing wrapper-based selection with and without prior filter-based ranking showed that filtering substantially improved robustness and consistency. Filter-based ranking effectively constrained the feature space, stabilized feature selection, and enhanced classification performance across all task configurations.

These results may support early PD assessment beyond specialist settings, as early symptoms are often first evaluated by general practitioners. Objective measures derived from standardized motor tasks could help facilitate earlier referral for neurological evaluation. Overall, this work moves beyond classification accuracy and emphasizes interpretable quantification of motor impairment and provides a more efficient, explainable, and clinically meaningful approach to characterizing early-stage PD.

## Data Availability

Due to ethical and privacy restrictions related to human participant data, the datasets used in this study are not publicly available.

## Acknowledgment

This work was supported by the DTU Discovery Grant and the DTU Alliance PhD Fund. The authors gratefully acknowledge the Movement Disorder and Pain Research Center at Rigshospitalet Glostrup, Denmark, for their invaluable contributions to study supervision and data collection.

